# The protein profiles of postoperative incision infection after plate internal fixation of calcaneal fractures

**DOI:** 10.1101/2023.06.28.23291984

**Authors:** Jun Wen, Zhong Wen

## Abstract

**Background:** Nowadays, the internal fixation has been an effective way for calcaneal fractures treatment. However, high risk of infection was found after the internal fixation, and the mechnism remains unclear.

**Objective:** In this work, we systematically preformed a comparative proteomic analysis between necrotic tissues and normal soft tissues aiming to find the molecular changes of the tissue during the fixation.

**Method:** The necrotic tissues (NTs) samples (n = 3) and the normal soft tissues control (NC) samples (n = 3) which was 2 -3 cm away from the NT were collected after the surgery. ALC-MS/MS analysis. A label free quantitation stragy was used to compare the proteome alterations followed by detailed bioinformatic analysis.

**Results:** A total of 902 and 1286 protein groups were quantified in the NT group and the NC group separately, with 233 proteins upregulated and 484 proteins downregulated in the NT group. Those differently expressed proteins were highly correlated with the metabolic pathways, especially those downregulated proteins in the necrotic tissue indicating an inacitive cell states in the nearby of the plate internal fixation. In addition, the detailed functiona analysis showed that the the upregutated proteins in necrotic tissue were highly enriched in the disease-related functions.

**Conclusion:** This alerted us to clean the wound in time and found a safer strategy for internal fixation. Altogether, the emerging understanding of the proteomic properties in the necrotic tissue will guide the development of new strategies for internal fixation of calcaneal fractures

## 1. INTRODUCTION

Calcaneal fractures constitute ∼ 60% of all tarsal fractures and 1% - 4% of fractures overall, making them the most commonly fractured bone of the tarsus.[1, 2] Though treatments of intra-articular calcaneal fractures remain controversial.[3] With the development of medical treatments, the fracture internal fixation has become an important therapy for calcaneal fractures.The Introduction section should include the background and aims of the research in a comprehensive manner.However, this approach can lead to a high risk of surgical site infection.[4, 5] Incision infection comprises one of the most frequent complications following a calcaneal fracture operation, involving deep structures such as bone, deep fascia, and implants, which may eventually lead to the formation of infection biofilms, osteomyelitis, scar formation, and even the need to remove the internal fixation.[5] Therefore, it is particularly necessary to found new strategy for safer treatment and prevent incision infection after the calcaneal fracture operation. Detailly, in the process of taking out the internal fixation material after the fracture internal fixation, we often see visible changes in color, texture and other aspects of soft tissues around the internal fixation. It is certain that some changes can’t be observed directly through our naked eyes. But no research has been conducted to dig out the protein changes of the surround tissues after infection. In order to have a more detailed understanding of the influence of metal internal fixation materials on the surrounding soft tissues, and the histopathological changes of the surrounding soft tissues under the influence of metal internal fixation after internal fixation, we performed a systematical and comparative proteomics analysis.

In our work, soft tissues with visible changes in color (blackening) and texture (softening) around the metal internal fixation were taken out for pathological examination during the surgical removal of internal fixation, and the soft tissues 2 – 3 cm away from the infection area were used as normal control. A robust and stable proteomic result were found in both necrotic tissues and normal tissues. Furtherly, according to integrative and comparative proteomic analyses, we found significantly dysregulated proteins in necrotic tissues were highly correlated with the inflammatory response, which alert us to develop new strategies and avoiding infection for internal fixation of calcaneal fractures.

## 2 MATERIALS AND METHOD

### 2.1 Chemicals and reagents

Sodium cholate (SDC), dithiothreitol (DTT), protease inhibitor cocktail and iodoacetamide (IAA) were purchased from Sigma-Aldrich (St. Louis, MO, USA). Acetonitrile (ACN, MS grade), trifluoroacetic acid (TFA, MS grade), formic acid (FA, MS grade) and the PierceTM BCA Protein Assay Kit were purchased from Thermo Scientific (St. Louis, MO, USA). Chromatographic-grade trypsin and Lys-C were purchased from Promega (Madison, WI, USA). Deionized water used in the experiments was prepared by the Milli-Q50SP system (Millipore, Milford, MA).

### 2.2 Patients and sample collection

The study was conducted following the guidelines of the Declaration of Helsinki, with the approval of the Research Ethics Committee from the Chinese Medicine Hospital of Xiangtan Country. Written informed consent was obtained from all patients or their legal representatives before they participated in the study. Samples were collected after surgery, and were washed three times with PBS. After that, tissues were immediately transferred to liquid nitrogen and collected into 2 mL cryogenic storage vials (Corning, New York, USA) and stored at − 80 °C for later use.

### 2.3 Sample preparation

One milligram of each sample was taken to grind into powder with liquid nitrogen in a pre-cooled mortar. Then the frozen sample powder of each group was transferred into 1.5 mL centrifuge tubes. And 1 mL of 1% sodium cholate (SDC) in 100 mM Tis-HCl buffer (pH 8.0) with 1X protease inhibitor cocktail were added for protein extraction. Samples were incubated at 95 °C for 30 min at 1000 rpm, followed by ultra-sonication for 10 min in ice bath (10 cycles; 30 sec ON/OFF). To remove the insoluble residues, each sample was centrifuged at 13000 g for 10 min at 4 °C. The resultant supernatants were successively transferred into new 1.5 mL centrifuge tubes and mixed with 10 mM dithiothreitol (DTT) for alkylation and bathed at 55 ℃ for 1 h. Then 30mM iodoacetamide (IAA) was added and rested for 30 minutes in the dark at room temperature. Protein concentration was determined using the PierceTM BCA Protein Assay Kit. An equal of 100 μ g protein for each sample was used for digestion. Then 2 µL of 1 µg/µl trypsin and 1µL of 1 µg/µl Lys-C were added to the samples for overnight digestion at 37 °C of 900 rpm. After digestion, 0.5% TFA was added to the sample for stop digestion and centrifuged at 4 °C for 10 min at 1300 g. Then the digests were desalted and dried using a SpeedVac centrifuge (Eppendorf, USA). The peptide mixture was then resuspended in 0.1% formic acid (FA) for LC-MS/MS analysis.

### 2.4 LC-MS/MS analysis

LC-MS/MS analyses were performed on the equipment of Orbitrap Exploris 480 mass spectrometer (Thermo Scientific) coupled with a Nano ACQUITY system (Waters Corporation). The ion source spray voltage is 2.2 kV, and the heating capillary was set at 320 ℃. Data dependence mode was adopted to automatically switch between MS and MS/MS. Full MS scans in Orbitrap mass analyzer were carried out with the scan range of 350 – 1600 m/z at a resolution of 120,000 (at 200 m/z). The automatic gain control (AGC) target value was set at 1x106, and the maximum injection time (IT) was 50 ms. High-energy collisional-trap dissociation (HCD) was applied to peptide fragmentation in the method of MS/MS model at the following 1.7 s, and the related data were obtained from scanning with the resolution of 30,000 by Orbitrap. The scanning range was automatically controlled by the mass charge ratio of parent ions, and the full scanning range was set from m/z = 110 to m/z = 2000. The minimum ionic strength values of MS/MS were set at 50,000 with a maximum IT of 45 ms. The AGC target value was set to 1.0 x 105, and parent ion mass tolerance was set at 1.6 Da. For analysis of LC-MS/MS data, charge states (+2, +3, +4) were taken into account. Ions selected for MS/MS were dynamically excluded for 30 seconds after fragmentation

### 2.5 Databased search

All raw data were analyzed by Peaks online (X build 1.7.2022-03-24_170623, Bioinformatics Solutions Inc.). The parameters were set as follows: MS 1 tolerance: 10 ppm; MS 2 tolerance: 0.02 Da; Searched Database: UniProt-human database (20,375 entries). A false discovery rate (FDR) of lower than 1% was used as the cutoff value for peptide, protein and propensity score matching (PSM) identification based on the target decoy strategy. Carbamidomethylation of cysteine was considered a fixed modification, and protein N-terminal acetylation, oxidation of methionine and deamidation of asparagine and glutamine were deemed to be variable modifications.ith 35% normalized collision energy.

### 2.6 Data analysis

Label-free quantification (LFQ) data were normalized by total ion chromatography (TIC). Quantified proteins with fold change > 2 or < 0.5 and P value < 0.05 were considered as differentially expressed proteins. In figures, experimental data are shown as standard error of mean.

The STRING[6] (https://www.string-db.org/) was utilized for functional enrichment and protein-protein interaction networks analysis. P values for the functional enrichments were calculated by hypergeometric test and corrected by Benjamini-Hochberg FDR method. And the MetaboAnalyst 5.0 (https://www.metaboanalyst.ca/)[7] was used for the statistical analysis (unsupervised cluster, heatmap, principal component analysis and volcano plot) of differentially expressed proteins. The proportional Venn diagrams and the GO analysis were analyzed by Bioinformatics online tool (http://www.bioinformatics.com.cn).

## 3. RESULTS AND DISCUSSION

The local pathology of the necrotic tissues (NTs): the periosteum around metal internal fixation (including part of the soft tissue near the periosteum) became dark and black, the soft tissue and blood vessels atrophied, the elasticity became poor or disappeared, partial soft tissue was denatured, and even the localized enveloping inflammatory changes appeared. Influence on healing: the above pathological changes of soft tissue leaded to local poor blood flow and low function of osteoblasts on periosteum, affecting the osteogenic response at the fracture site and delaying fracture healing.

To systematically analyze the protein dynamics in NTs after internal fixation, we conducted the proteomic analysis of NT compared with soft tissues 2 – 3 cm away from the NT as normal control (NC). Though, similar missed cleavages were in both groups, the proteomic analysis showed a lower identification of NT compared with NC (Figure 1A and B). In detail, an average of 1125 proteins, 8030 peptides and 12214 PSMs were identified in NC group with three replicates, while only an average of 742 proteins, 8030 peptides and 7483 PSMs were found in NT group, indicating the necrotic tissue has denatured. Furtherly, high coverage of quantified proteins were found in each group, with a total of 902 LFQ proteins in NT group (n = 3) and 1286 LFQ proteins in NC group (Figure 2C and D). What’ s more, low correlations of the total quantified protein and peptides in two groups, with only 32.45% and 14.16% were co-quantified in proteins and peptides, respectively (Figure 1E and F), indicating the necrotic tissues had significantly changes in their proteome component.

**Figure 1.**
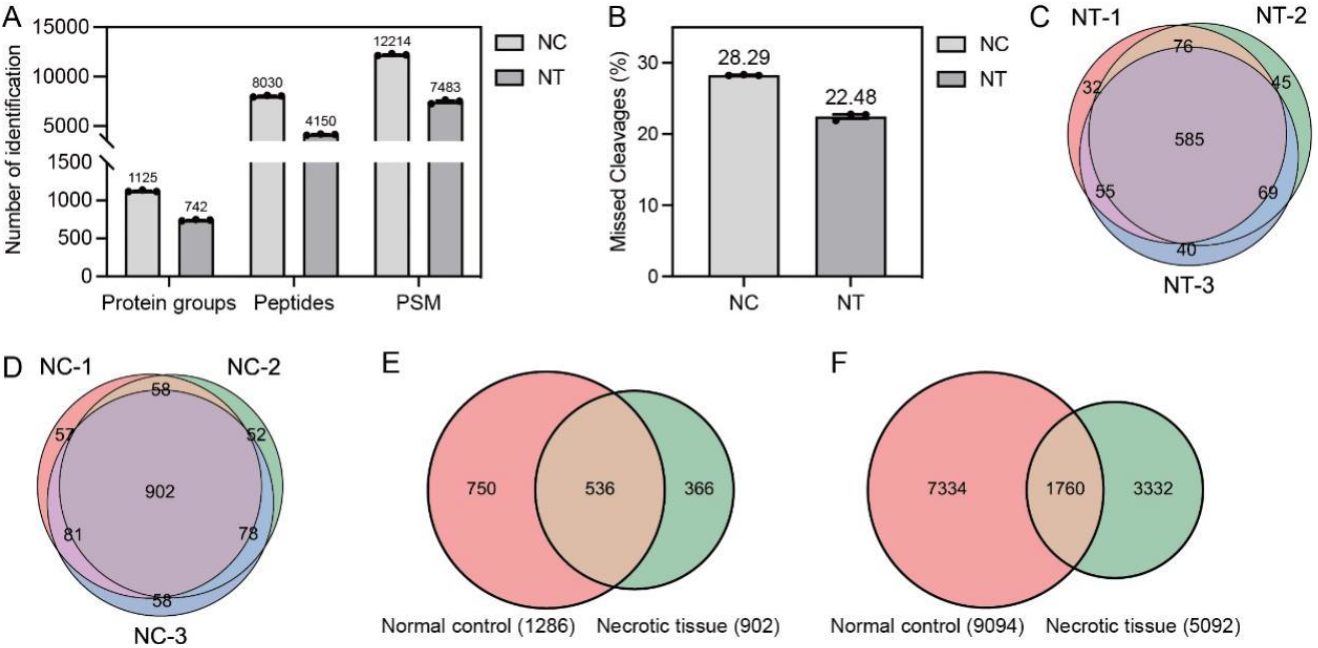
Proteomic analysis of necrotic tissue (NT) versus normal control (NC). (A) Number of identified protein groups, peptides and PSM in two groups (mean ± SEM, n = 3). Values above columns indicate average numbers of identification. (B) Percentage of missed cleavages in two groups. (C and D) Proportional Venn diagram of total quantified proteins (C) and peptides (D) in two groups.

**Figure 2.**
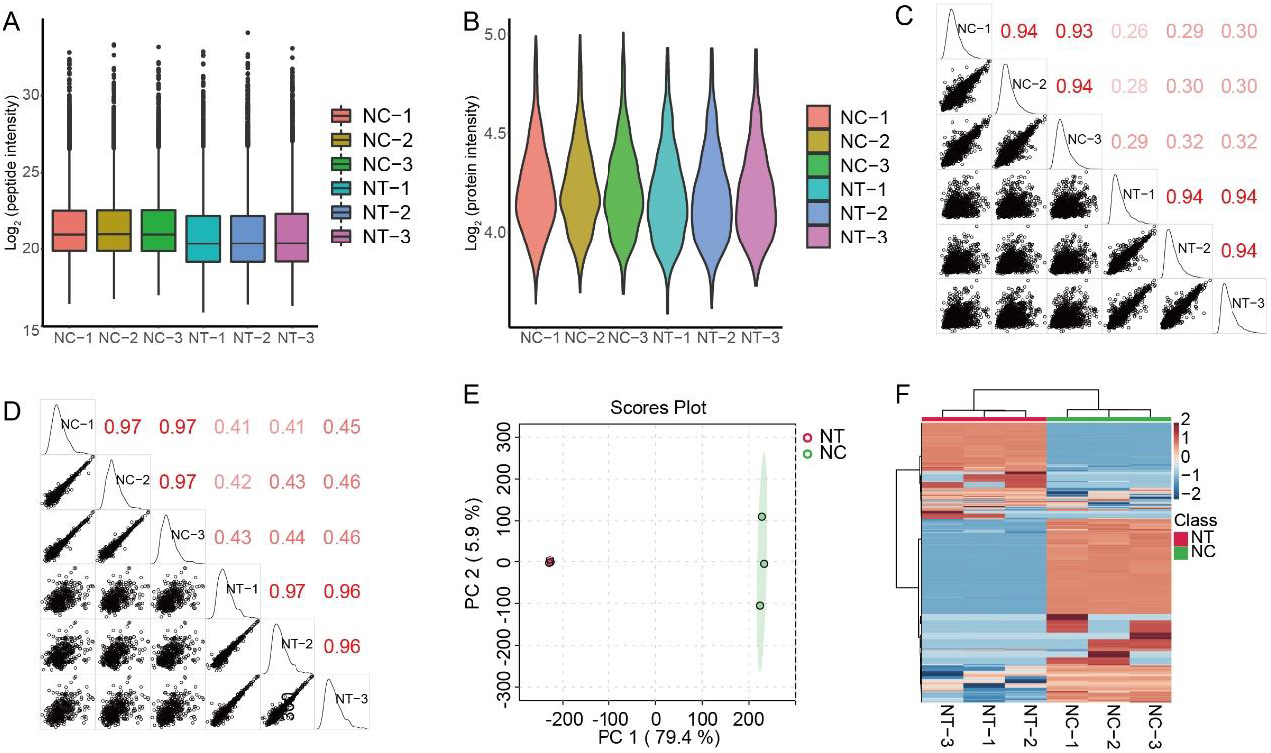
Integrative analysis of two groups. (A) Boxplot of peptide intensity distributions. (B) Violin plot of peptide intensity in each group. Box limits indicate the 25th and 75th percentiles as determined by R software; whiskers extend 1.5 times the interquartile range from the 25th and 75th percentiles; polygons represent density estimates of data and extend to extreme values. (C and D) Pearson correlation of quantified peptides and proteins in NC and NT groups (n = 3). (E) Principal component analysis (PCA) of NT (n = 14) and NC (n = 13) samples based on LFQ proteins. (F) Multigroup heatmap with dendrogram of LFQ proteins across 6 samples. Ward’s method was performed for clustering, and the Euclidean distance was used for distance measurement.

Additionally, stable peptide and protein intensities were detected between necrotic tissues and soft tissues (Figure 2A and B), suggesting a reliable proteomic analysis that allowed us for further analysis. Meanwhile, high Pearson correlations were found in both groups, with an average of 0.93 and 0.94 of LFQ peptides in NC and NT group separately, as well as 0.97 and 0.96 of quantified proteins in each group (Figure 2C and D). And the principal component analysis (PCA) effectively distinguished two types of tissues based on component 1 (79.4%) and component 2 (5.9%) (Figure 2E). In addition, the unsupervised cluster showed two separate entities of NC and NT groups, indicating a high difference between necrotic tissues and normal tissues (Figure 2F).

Therefore, we conducted the difference analysis based on volcano plot, resulting 717 differently expressed proteins (DEPs), of which 233 proteins were upregulated and 484 proteins were downregulated (Figure 3A, adjust P < 0.05). For instance, the S100 protein family such as S100B and S100AB were distinctly changed in necrotic tissues, of which the S100B was downregulated while the S100AB was upregulated (Figure 3B). The S100 proteins show cell-specific expression patterns and participate in many cell process including innate and adaptive immune responses, cell migration and chemotaxis, tissue development and repair, and leukocyte and tumor cell invasion.[8] The over expressed S100AB protein, a heterodimeric EF-hand Ca2+ binding protein, has been widely explored as a key protein between inflammatory processes and several cancerous pathogens. [9, 10] Besides, the decorin (DCN) protein, the prototype member of the small leucine-rich proteoglycans, was regarded as a tumor regulator by downregulating the activity of several receptors involved in cell growth and survival. [11] Another representatively upregulated protein was closely related to tumor genesis, the keratin17 (KRT17). KRT17 functions as a tumor promoter and regulates proliferation, migration and invasion in many malignancies. [12, 13] Taken together, those DEPs were highly correlated with tumor genesis, indicating a high risk of the postoperative incision infection after plate internal fixation of calcaneal fractures.

**Figure 3.**
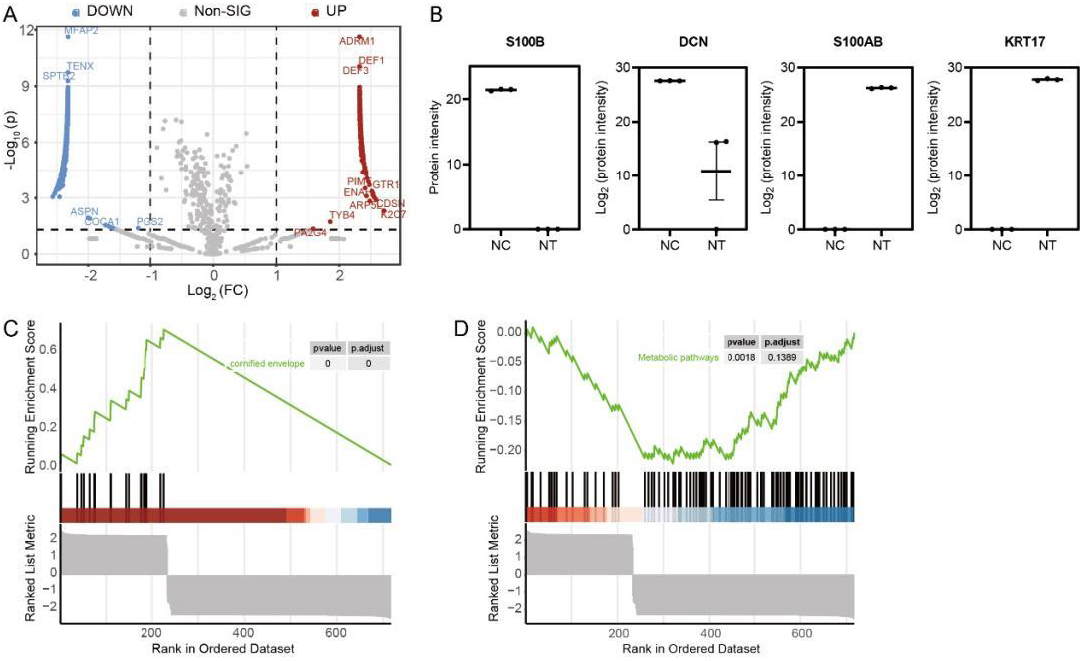
(A) Volcano plot of the statistical significance between two groups (two-tailed Student’s t-test, adjust P < 0.05 and fold change > 2 or < 0.5). (C and D) The gene set enrichment analysis (GSEA) of differently expressed proteins in NT compared to NC based on the GO (C) and KEGG (D) terms (Benjamini− Hochberg FDR method, adjusted P < 0.01).

Ontology (GO) analysis – cornified envelope, with aIn addition, the gene set enrichment analysis (GSEA) of DEPs showed one highly variated term based on the Genetendency to rise in the necrotic tissue and fall in the normal soft tissue (Figure 3C). The cornification is a symbol of cell death,[14] indicating a variety of cell necrosis-related proteins were detected in the center of plate internal fixation.

Meanwhile, the KEGG analysis revealed the significantly changed pathway was metabolic pathway, of which downregulated in the necrotic tissue and upregulated in the normal tissue, also revealing a variety of cell deaths in the nearby of the plate internal fixation (Figure 3D).

Furthermore, we performed a detailed functions enrichment of those DEPs, to dig out some new findings to explain the necrosis process and found better treatments. The GO analyses were conducted based on the biological process (BP), cellular component (CC) and molecular function (MF), separately. And the top 10 annotations of each analysis were showed (Figure 4A). Surprisingly, GO-BP was focused on cell death-related terms such as extracellular matrix organization, cornification, neutrophil degranulation, this was consistent with the GSEA results above (Figure 4A and B). In detail, the collagen family (e.g., COL10A1, COL12A1, COL15A1), the major component of extracellular matrix, was found in BP analysis, which was also regarded as the major component of the tumor microenvironment and participates in cancer fibrosis.[15] Similarly, the lamin protein family (e.g., LAMB1, LAMA5, LAMC1), a basement membrane glycoprotein, also showed distinct variations in the NT versus NC. The lamin protein family has been implicated in a number of stages in tumour invasion and metastasis.[16] In concordance with the previous conclusion, the S100 protein family (e.g., S100A7, S100A8, S100A9) and the keratin family (e.g., KRT5, KRT7, KRT14) were mostly upregulated in the NT group compared with NC.

**Figure 4.**
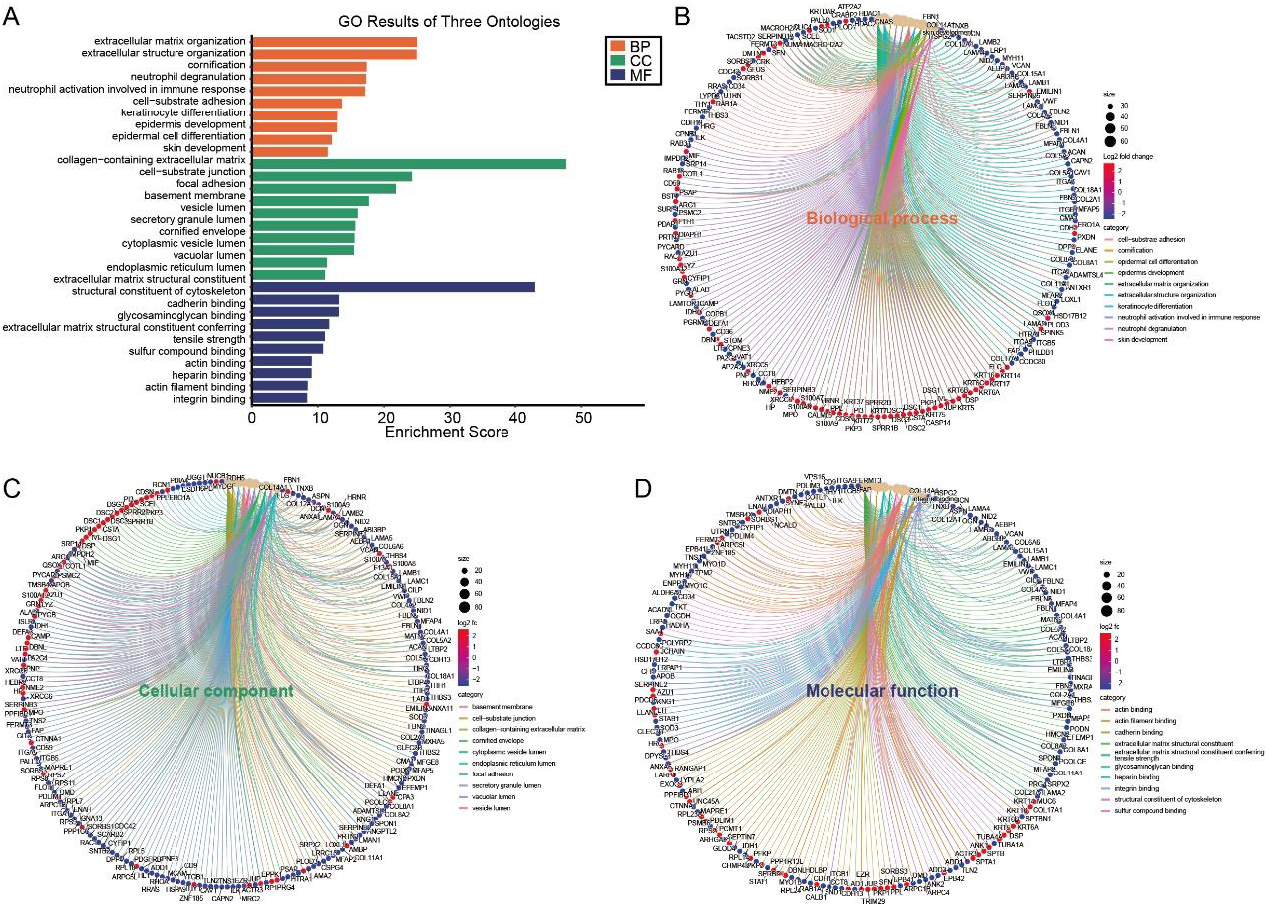
Functions enrichment of differently expressed proteins (DEPs). (A) The Gene Ontology (GO) analysis of DEPs. The top 10 annotations of biological process (BP), cellular component (CC) and molecular function (MF) are shown (Benjamini− Hochberg FDR method, adjusted P < 0.01). (B-D) GO chord showing the interactions between up and downregulated proteins in BP (B), CC (C) and MF (D) analysis.

**Figure 5.**
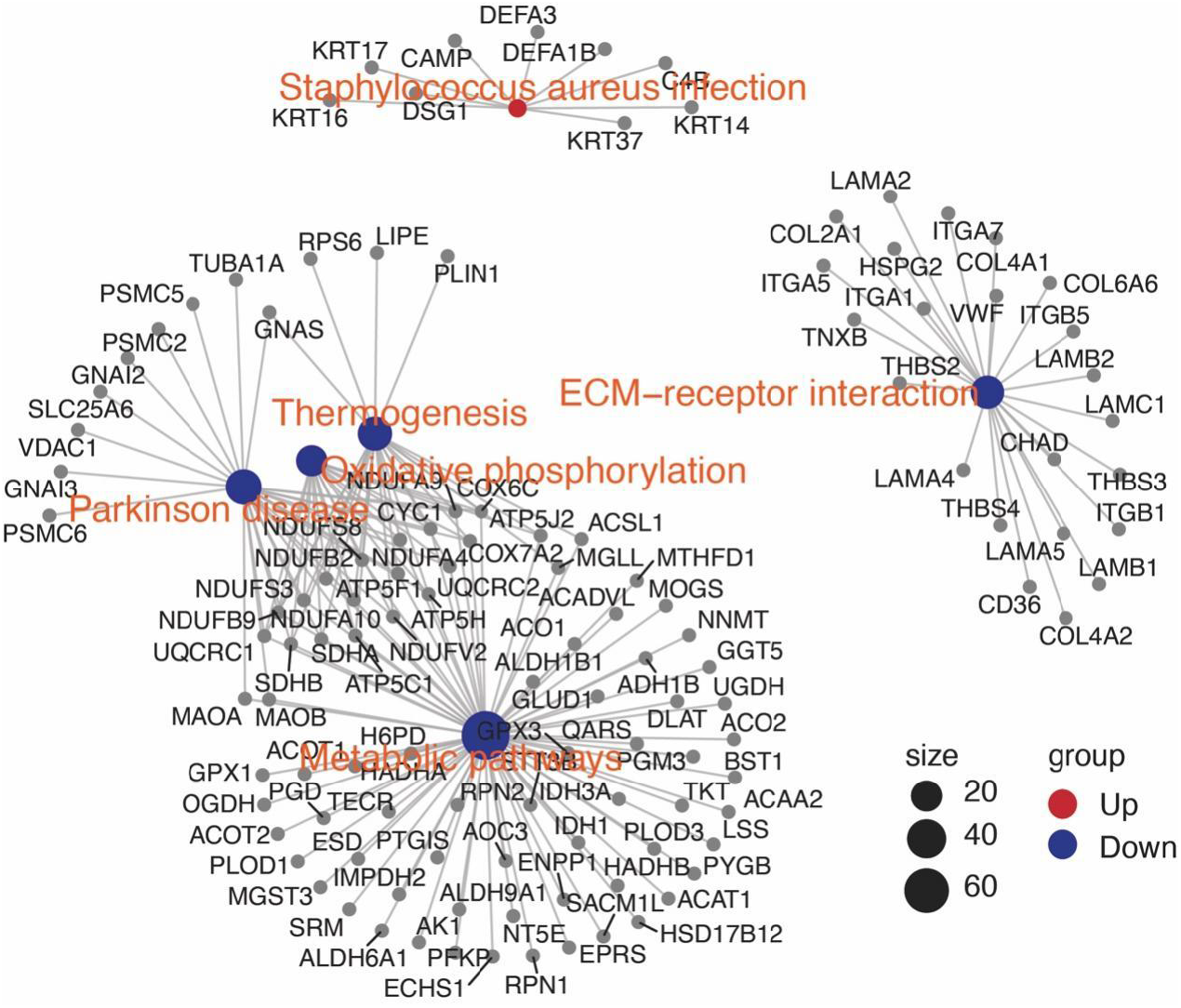
Protein network analysis showing high connection of up and downregulated proteins in necrotic tissue (the Benjamini− Hochberg FDR method, adjusted p < 0.05).

Apart from the GO-BP result, the GO-CC was mainly enriched on cell− substrate junction, vesicle lumen and basement membrane (Figure 4A and C). In cell− substrate junction, a series of desmogleins (e.g., DSG1, DSG3) and desmocollins (e.g., DSC1, DSC2, DSC3) were upregulated in the necrosis part, which were also found to be dysregulated in various cancers, revealing a tight correlation between tumorigenesis and tissue necrosis.[17, 18] And for the vesicle lumen, indicated the decreased secretion of collagens (e.g., COL6A6, COL12A1, COL15A) for cell communications and cell connections. And the basement membrane, composed with a variety of dysregulated collagens, laminins and integrins (e.g., ITGA5, ITGB5, ITGA1), was closely related to tumor metastasis.[19]

Moreover, a series of cell binding-correlated terms was found in the GO-MF analysis, including integrin binding, actin binding, cadherin binding and surprisingly, the glycosaminoglycan binding (Figure 4A and D). The integrins and cadherins was consistent with the GO-BP and GO-CC analyses that the necrotic part triggered a series of inflammatory reactions. While the glycosaminoglycan binding gave us a hint that the necrotic tissues existed a large number of glycoproteins (e.g., FBN1, TNXB, ANXA6), which was also present in cancer and reported to correlate with clinical prognosis in several malignant neoplasms.[20] In conclusion, the detailed GO analyses suggested that the necrotic tissue showed a similar protein pattern with tumors, which alerted us to clean the wound in time and found a safer material for internal fixation of calcaneal fractures.

Additionally, we also analyzed the enriched KEGG pathways of significantly upregulated or downregulated proteins individually. For upregulated proteins in NT, only one pathway was found to be statistically correlated, the staphylococcus aureus infection, of which the keratin family was detected and other novel discovered proteins. For instance, the complement C4-B protein (C4B) participated in complement process and regarded as novel anti-tumor strategy.[21] And the cathelicidin antimicrobial peptide (CAMP) was also found to be upregulated in cartilage tissue, which induced chondrocyte apoptosis.[22] Surprisingly, the increased expressions of neutrophil And for the KEGG analysis of downregulated proteins, 63 pathways were found that mainly focused on cell metabolism and disease-related processes. We chose five representative pathways for further analysis, including ECM-receptor interaction, thermogenesis, Parkinson disease, oxidative phosphorylation and metabolic pathways. Importantly, the thermogenesis, Parkinson disease and oxidative phosphorylation pathways were highly correlated with metabolic pathways, indicating a close relationship between tissue necrosis and energy metabolism. Another significantly downregulated pathway was ECM-receptor interaction, also representing the cell death in the necrotic part. Together, based on the KEGG analyses, we found the necrotic tissues after internal fixation of calcaneal fractures contain large amounts of cell apoptosis-related dysregulated proteins which may lead to inflammatory.

## CONCLUSION

In this work, we systematically preformed a comparative proteomic analysis between necrotic tissues and normal soft tissues. A comparable and robust result was obtained that a total of 902 LFQ proteins in NT group (n = 3) and 1286 LFQ proteins in NC group. By integrative analyses, the distinct protein profiles between two groups were found that allow us for further difference analysis. The volcano plot generated 233 upregulated proteins and 484 downregulated proteins. Those DEPs were highly correlated to cornified envelop and metabolic pathway, reveling the cell death and low energy metabolism in the necrotic tissue. In addition, the detailed GO and KEGG analyses showed that the necrotic tissue contains a variety of disease-related dysregulated proteins. This alerted us to clean the wound in time and found a safer strategy for internal fixation. To sum up, the emerging understanding of the proteomic properties in the necrotic tissue will guide the development of new strategies for internal fixation of calcaneal fractures.

## Data Availability

All data produced in the present study are available upon reasonable request to the authors

http://proteomecentral.proteomexchange.org

## ETHICS APPROVAL AND CONSENT TO PARTICIPATE

The study was conducted following the guidelines of the Declaration of Helsinki, with the approval of the Research Ethics Committee from the Chinese Medicine Hospital of Xiangtan Country. Written informed consent was obtained from all patients or their legal representatives before they participated in the study.

## SUPPORTIVE/SUPPLEMENTARY MATERIAL (IF ANY)

The mass spectrometry proteomics data have been deposited to the ProteomeXchange Consortium (http://proteomecentral.proteomexchange.org) via the iProX partner repository[23, 24] with the dataset identifier PXD041483.

